# Rare genetic variants influence participation in cognitive components of the UK Biobank

**DOI:** 10.64898/2026.10.08.26365060

**Authors:** Eilidh Fenner, Peter Holmans, Michael C O’Donovan, Michael J Owen, Elliott Rees, James T R Walters

## Abstract

Population-level biobanks are transforming health and genomic research however they are all subject to recruitment and participation biases, the impacts of which are not fully understood. The UK Biobank (UKBB) is one such cohort; it is widely used for health-related research, but its participants are not representative of the UK general population. In addition, those who complete optional components of UKBB differ from those who do not in terms of health-related exposures and outcomes. Participation in UKBB has a partly genetic basis, but the extent to which rare variants influence participation in optional study components, and how this might impact findings of rare variant association studies, is unknown.

We analysed exome sequencing and array data from 398,149 UKBB participants to examine rare variant contributions to UKBB participation bias, focusing on cognitive phenotypes. We found that classes of rare variation previously shown to contribute to cognition were enriched among participants who completed fewer cognitive tests. We then compared the effects of rare variants on cognition itself between groups of participants defined by whether they completed all online cognitive tests. Rare variants had significantly weaker effects on cognition in individuals who completed all online cognitive tests than in those who did not.

These findings demonstrate that participation is a non-random phenotype that is influenced by rare variants, and that individuals who engage in optional UKBB components are not representative of the broader cohort. We show how participation bias can reduce the power of rare variant association studies, potentially masking true associations. This bias should therefore be carefully considered when interpreting results derived from phenotypes with substantial missingness.

## Introduction

Participation bias is a well-recognised concern for studies using large-scale cohorts such as the UK Biobank (UKBB). Genetic factors can influence whether an individual takes part in a study at all, and whether they participate in optional and follow-up study components. Genetic studies relying on optional study components with some missingness may therefore be biased. While the influence of common genetic variants on study participation is established^1–4^, much less is known about the contribution of rare variants, including rare coding variants (RCVs) and rare copy number variants (CNVs).

UKBB includes numerous optional components, such as repeat assessment centre visits, online questionnaires, and cognitive testing, recorded over up to 20 years. The availability of genetic data in this cohort offers an opportunity to study how genetic variants may influence participation across optional or longitudinal data capture episodes. Previous studies have highlighted the role of common alleles in participation in optional components of UKBB, and have shown that genetic factors influencing participation are correlated with those influencing demographic and health-related factors^1,5,6^. Here, we aim to examine whether rare variants similarly influence participation in optional components of UKBB and assess the impact of any such biases on interpretation of results that use optional phenotypic outcomes.

In this study, we focus mainly on participation in cognitive components because cognitive ability has established associations with rare genetic variants; for example, a higher burden of damaging RCVs^7–9^ and rare CNVs^10,11^ is associated with lower scores on various measures of cognitive ability in UKBB. Cognitive ability is also measured repeatedly, across many optional assessments, in UKBB, allowing for the derivation of phenotypes reflecting the level of participation in cognitive testing. This study therefore tests whether the types of rare variants that impact cognitive ability also influence participation in cognitive testing in UKBB. If this is the case, it would imply that UKBB participants who complete more cognitive tests are less likely to carry damaging rare variants, potentially biasing effect sizes and reducing power to detect associations between these variants and cognition itself.

Participation bias may be particularly relevant for studies of genetic factors that increase liability for neurodevelopmental and psychiatric disorders (NPDs), such as schizophrenia^3^ and attention deficit hyperactivity disorder (ADHD)^12^. For example, higher polygenic scores for schizophrenia and ADHD are associated with loss to follow-up^3,12^, suggesting individuals with higher liability for NPDs may be underrepresented in optional components of population-level cohorts. As many NPDs are associated with cognitive impairment, genetic liability for these disorders likely contributes to both lower cognitive ability and to reduced participation in cognitive testing, resulting in underrepresentation of those with high genetic liability for NPDs in optional cognitive assessments.

Participation bias has the potential to reduce power to detect genetic associations, distort estimates of these associations, and limit the generalisability of findings to less biased or clinical cohorts^13–17^. Several methods have been developed to mitigate these effects, many of which involve reweighting participants based on their representativeness of a target population^4,14,15,18,19^. Here, we build on this work by exploring how participation bias impacts rare variant association studies that rely on optional study components, and by evaluating whether reweighting approaches can reduce the impact of this bias.

To our knowledge, the current study provides the first systematic assessment of rare variants and participation in optional UKBB study components. We show that individuals who carry damaging RCVs and rare CNVs are less likely to participate in optional cognitive assessments. Importantly, this finding is not fully explained by reduced cognitive ability in carriers of damaging RCVs and rare CNVs, suggesting a broader impact of rare variation on study engagement. We then present novel evidence demonstrating that participation bias can distort, or even mask, rare variant-phenotype associations, highlighting the need to interpret findings of rare variant association studies (particularly null results) in the context of participation biases. Overall, this work underscores the need for careful consideration of participation-related factors in genetic studies of population-level cohorts.

## Methods

Between 2006 and 2010, UKBB recruited approximately 500,000 participants from the UK through NHS registers, with no exclusion criteria apart from reasonable access to an assessment centre. Participants were aged 40-69 at recruitment and 54% were female. All participants gave consent for their data to be used by UKBB projects and agreed to being followed up. Ethical approval was granted to UKBB by several committees, as it spans England, Scotland, and Wales: The Northwest Multi-Centre Research Ethics Committee; The Patient Information Advisory Group, which has since been replaced by the National Information Governance Board for Health and Social Care; and The Community Health Index Advisory Group. The research presented in this study was conducted using UKBB under Application Number 13310.

## Genetic data

### Whole exome sequencing data

Whole exome sequencing (WES) data were generated using the IDT xGen Exome Research Panel v1.0 exome capture kit and Illumina NovaSeq 6000 instruments. S2 flow cells were used for the initial 50k sequencing release, and S4 flow cells were used for all other samples. Details on sequencing protocol can be found in the primary UKBB publication^20^. These data, available for 469,385 participants, were accessed and analysed using UKBB Research Analysis Platform (RAP), and processed as described in our previous publication^7^.

### Quality control

WES data were quality controlled using Hail (version 0.2.60^21^) and R (using RStudio v2.2.1) on the RAP. Genotype-level quality control (QC) excluded genotypes with depth ≤ 10x, genotype quality ≤ 30, or unusual allele balance (homozygous reference with an allele balance > 0.1, homozygous alternate with an allele balance < 0.9, or heterozygous genotypes with an allele balance < 0.25 or > 0.75). Variants were excluded if they: were labelled as ‘mono-allelic’^22^; had > 6 alternative alleles; had no alternative alleles after applying genotype QC; were on the X or Y chromosome (as our analysis focused on autosomes); had low mean genotype quality (≤ 40); had low call rate (≤ 0.9); or overlapped a low-complexity region. The original dataset included 22,854,479 variants, of which 19,724,617 remained following genotype and variant QC.

Sample level QC (further detail available in ^7^) then excluded individuals with: low sequencing call rate (< 0.8, n = 234); discordant self-reported sex, imputed sex, and/or Y chromosome depth (n = 210); or insufficient data for inferring ancestries or calling CNVs (lacking or low-quality array data, n = 1,518). 69,274 related individuals (third degree relatives or closer, kinship co-efficient > 0.0442^23,24^) were also excluded, retaining 398,149 unrelated individuals with high quality genetic data for analysis.

### Rare coding variant annotation

RCVs (single-nucleotide variants and insertions and deletions < 50 base-pairs in length) were annotated in Hail using Ensembl’s Variant Effect Predictor (VEP)^25^, and defined as variants with a minor allele count ≤ 5 within the post-QC sample (equating to a minor allele frequency < 6.25 x 10^-6^), a definition similar to that of other UKBB RCV studies^9,26^. Damaging RCVs were defined as rare protein-truncating variants (PTVs; splice acceptor/donor, stop-gain or frameshift variants) annotated as high confidence for causing loss of protein function by LoFTEE^27,28^, and deleterious missense variants with REVEL^29^ scores > 0.75. We also examined rare synonymous variants as a negative control.

### Gene sets

We focused on gene sets in which RCVs have been shown to be associated with cognitive ability, as we aimed to investigate whether these same genes are important in participation in cognitive testing. We therefore analysed damaging RCVs in the following autosomal protein-coding gene sets:

#### Loss-of-function intolerant (LoFi) genes

3,162 genes under selective constraint, defined as genes with a GnomAD pLI score^30^ ≥ 0.9. RCVs in this set have been associated with various measures of cognitive ability in UKBB^7,9,31^.

#### Neurodevelopmental and psychiatric disorder (NPD) associated genes

770 genes, including (with some overlap): (i) 655 genes in which rare variants are known to cause monogenic severe developmental disorders, taken from the developmental disorders genotype to phenotype (DDG2P) (v5.1) panel^32,33^; (ii) 29 genes enriched for RCVs in schizophrenia at a false discovery rate of < 5% in a recent large-scale sequencing study of schizophrenia (SCHEMA^34^); and (iii) 101 credibly causal genes implicated in schizophrenia by the most recent Psychiatric Genomics Consortium genome-wide association study of schizophrenia. RCVs in each of these sets individually have been associated with lower cognitive ability in UKBB^7,8^.

Table 1 presents the number of variants within each of these sets, and the distribution of these variants across individuals.

**Table 1.**
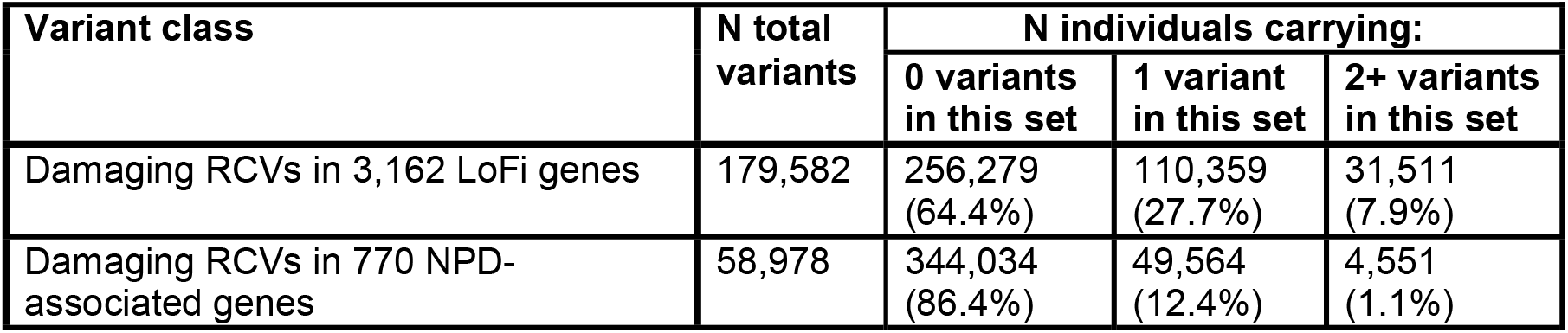
Number of variants of RCVs in two different gene sets carried in this cohort (n = 398,149), and number of people carrying different numbers of these variants. RCV = rare coding variant; LoFi = loss-of-function intolerant; NPD = neurodevelopmental and psychiatric disorder.

### Array data

Participants were genotyped using the Affymetrix Axiom UKBB array (∼450,000 individuals) and the UK BiLEVE array (∼50,000 individuals).

#### Copy number variant calling

Array data were used to identify carriers of NPD-associated CNVs, which have been shown to be associated with lower cognitive ability in UKBB^10,36^. These CNVs were called as described in our previous publications^10,11^. 4,311 NPD-associated CNVs were observed in this cohort, with 4,235 participants carrying only one of these CNVs (1.06%), and 38 participants (0.01%) carrying 2 or more. The majority of our cohort (n = 393,876; 98.9%) did not carry any NPD-associated CNV.

#### Genetic ancestry inference

Analyses for inferring genetic ancestry utilised UKBB array data, biogeographical categories based on a standardised system^37^ and our previously published methods^7,38,39^. In short, UKBB participants were projected into principal components (PCs) derived from the 1000 Genomes Project (1KGP) Phase 3 reference panel^40^. Genetic ancestry probabilities were estimated using a model based on Fisher’s Linear Discriminant Analysis^41^ (which was trained on known ancestries of the 1KGP reference samples, and then applied to the UKBB dataset) and genetic ancestry labels were assigned based on these probabilities. Individuals were assigned a 1KGP-like genetic ancestry label if any ancestry probability was > 0.8 and otherwise were assigned an ‘Admixed’ label. These labels reflect only genetic similarity to the 1KGP reference panel^40^ and do not represent biologically distinct categories.

Given substantial differences in rare variant burden across 1KGP-like genetic ancestry groups (Supplementary Table 1 and ^7^), we analysed individuals in each 1KGP-like genetically inferred ancestry group of sufficient sample size (> 5,000 individuals) separately.

## Phenotypic data

### Participation in cognitive testing

Phenotypic data were collected from UKBB participants at multiple timepoints, in person and online. Between 2006 and 2010, all participants attended an initial assessment centre (henceforth referred to as baseline) where data were collected via touchscreen questionnaires, interviews, and sample collection. Since 2012, participants have been recontacted via email and post to participate in optional components of UKBB (see Figure 1 for detail on cognitive data collection).

**Figure 1.**
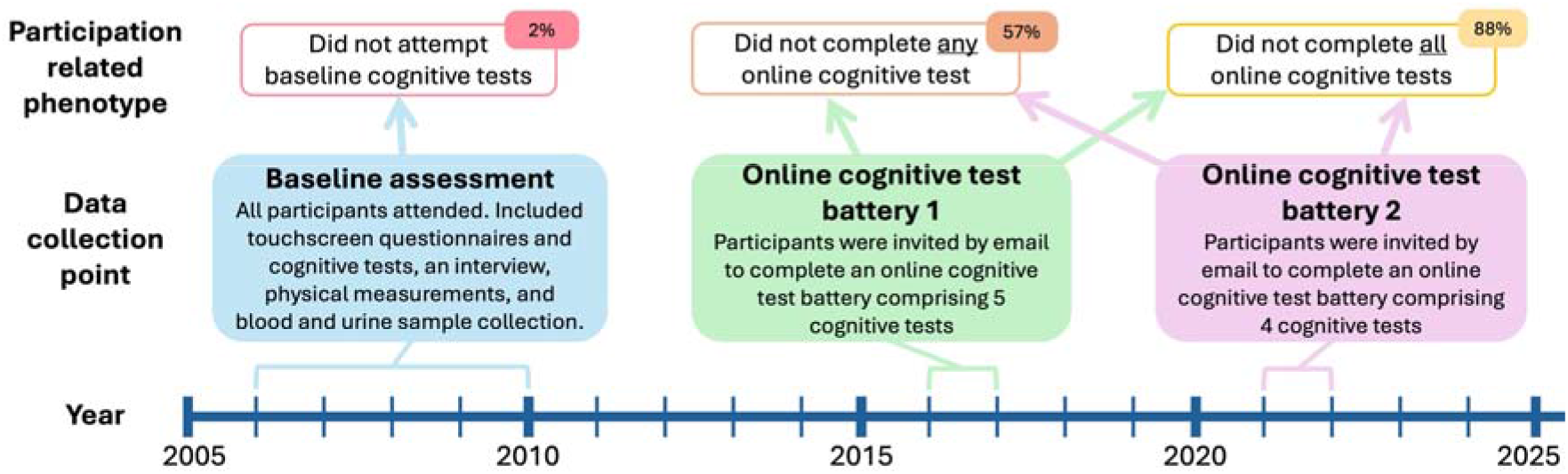
Timeline of cognitive data collection points in UKBB including detail on the data collected. Participation-related phenotypes are listed in the top row of this diagram, with arrows to show which data collection point these are derived from.

We derived three binary phenotypes reflecting participation in cognitive testing in UKBB, described in detail below (and in Supplementary Table 2):

#### Did not attempt baseline cognitive tests

During baseline assessment centres, participants were asked to complete cognitive tests on a touchscreen device by being presented with a screen stating *‘Now we would like to check your memory and reaction times by getting you to play some short games’*. They could select either *‘Begin games’*, which took them to the baseline cognitive tests (97.9%), or *‘I am unable to try this’*, which meant they did not undertake any of the baseline cognitive tests (2.0%) (UKBB Field 62).

#### Did not complete <u>any</u> online cognitive test

UKBB participants were invited via email to complete two online ‘Healthy Minds’ assessments, once between 2014-2015, and then again between 2021-2022 (Figure 1). These assessments involved answering questions about mood and completing either 4 or 5 cognitive tests. We examined whether someone completed any of the 9 online cognitive tests from across the two assessments. 57.2% of participants did not complete <u>any</u> online cognitive test.

#### Did not complete <u>all</u> online cognitive tests

Here, we similarly used data from the online ‘Healthy Minds’ assessments, and examined whether someone completed all 9 online cognitive tests (and thus had ‘complete’ online cognitive test data). 87.6% of participants did not have test scores recorded for <u>all</u> cognitive tests from the online assessments.

Many participants were never invited to complete online cognitive testing (usually due to not providing a working email address) and we therefore ran sensitivity analyses controlling for invitation to online testing. We also examined three additional broad and non-cognitive participation phenotypes (lost to follow-up (30.1%), no email access (40.3%), did not complete mental health questionnaire (68.3%)), described in Supplementary Materials 1.1.

### Measures of cognitive ability

Two measures of cognitive ability were used in this study: reaction time (RT), a test reflecting processing speed which was available for 392,969 participants at baseline (98.7%); and fluid intelligence (FI, often referred to as verbal-numerical reasoning), a test measuring reasoning ability, which was available for 250,915 participants (63.0%) from across different instances (Supplementary Table 3). Supplementary Materials 1.2 presents further detail on these phenotypes.

### Reweighting

To account for the impact of participation bias on the associations of rare variants with phenotypes, we derived stabilised inverse probability weights based on the estimated probability of completing <u>all</u> online cognitive tests (and thus having ‘complete’ cognitive test data). We selected demographic variables for use as predictors of participation, selecting phenotypes that: have been used in previous reweighting studies of UKBB^15,18,19,42^; were recorded at baseline assessment; and were available for > 99% of the cohort. Phenotypes included in reweighting were age, sex, ethnicity, country of birth, socioeconomic status, relationship status, and early-life factors, and are described in full in Supplementary Table 4. The processing of these variables is described in Supplementary Materials 1.3. Participation (coded as 1 = completed <u>all</u> online cognitive tests, 0 = did not complete <u>all</u> online cognitive tests) was modelled as the dependent variable in a logistic regression, with these demographic variables included as predictors.

In total, 392,812 individuals had complete data for all reweighting phenotypes and were alive at the time of invitation to all cognitive tests (98.7% of the cohort). For each participant, we estimated the probability of participation from the logistic regression model, and derived stabilised inverse probability weights by scaling the inverse probabilities by the overall participation rate, similarly to other studies^18,42^. Weights were capped at 4SDs above the mean for 3,142 participants with weights outside of this.

## Statistical analyses

### Non-participation and rare variant burden

Logistic regression models were used to investigate the association between measures of participation (dependent variable) with burden of rare variants (independent variable), tested separately for each measure of participation (participation in baseline cognitive tests; completed <u>any</u> online cognitive test; completed <u>all</u> online cognitive tests), and each of the three rare variant classes (damaging RCVs in LoFi genes, damaging RCVs in NPD-associated genes, and rare NPD-associated CNVs). Given the low frequency of carriers of some rare variant classes (e.g. NPD-associated CNVs), we used bias-reduced logistic regression (via the brglm2^43,44^ R package), which applies Firth-type penalised likelihood estimation. All reported regression p-values are two-sided. Our primary analysis tested these three rare variant classes across three phenotypes. To account for multiple testing in this primary analysis (3 x 3 = 9 tests), we applied a Bonferroni correction resulting in a significance threshold of 0.05/9 = 0.0056.

In these models, we included covariates to control for population stratification, including: ancestry probabilities (for ancestries for which there were at least five people with a non-zero probability), and excluding 1KGP-EUR-like probability (inclusion of all ancestry probabilities induces co-linearity); and within-ancestry PCs 1-10 (e.g. for analysis of the 1KGP-EUR-like population, the first 10 PCs derived from just this population were included as covariates). In addition, we covaried for assessment centre attended. For analyses of damaging RCVs, we also included burden of synonymous variants (except when investigating synonymous variant burden) and sequencing batch as covariates. For analyses of CNVs, we included array batch as a covariate. As participation (and factors influencing it) may vary non-linearly across age, and as this could differ by sex^45^, we also covaried for: sex; standardised age; standardised age squared; and interactions between sex and standardised age, and between sex and standardised age squared.

We ran exploratory analyses to expand on our primary findings, using the same statistical approach described for the primary analysis and the same significance threshold of 0.0056. First, we tested whether our findings were specific to participation in cognitive testing, or whether they applied to participation more generally, by analysing three additional, broader participation-related phenotypes (described in Supplementary Materials 1.1). We then added additional covariates to our primary analyses to examine whether our findings were driven by any of the following factors: (i) invitation to cognitive testing; (ii) NPD diagnoses; and (iii) cognitive ability, as we hypothesised that those with higher cognitive ability may be more likely to choose to participate in cognitive testing. Further detail on the processing of these additional covariates is presented in Supplementary Materials 1.4. To compare associations with and without these additional covariates, we performed z-tests using the equation

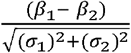

where: *_β_*_1_ is the beta and *_σ_*_1_ is the standard error of association with the usual covariates; and *_β_*_2_ is the beta and *σ*_2_ is the standard error of association in the model with an additional covariate for invitation, diagnosis, or cognitive test score.

#### Impact of non-participation on rare variant associations with cognitive ability

To investigate the impact of participation bias on observed effects of damaging RCVs and rare CNVs on cognitive ability in UKBB, we ran linear regressions testing for associations between cognitive ability (RT and FI) and burden of damaging RCVs and rare CNVs in two subsets of the cohort: those who had completed <u>all</u> online cognitive tests (i.e. had ‘complete’ cognitive test data, n = 49,391); and those who had not (n = 348,758). All models included the same covariates described in the primary analyses. We then compared association effect sizes between groups using z-tests from the equation previously described. We ran these z-tests across the three classes of rare variants and across two cognitive measures (3 x 2 = 6 z-tests). We therefore applied a Bonferroni correction to z-test p-values resulting in a threshold of 0.05/6 = 0.0083.

Reweighting analyses were run as described above, with stabilised inverse probability weights applied to the linear regressions. We also fit regression models in the full sample including an interaction term between rare variant burden and participation (whether someone had completed <u>all</u> online cognitive tests) to confirm that differences observed in the stratified analyses were consistent across analytical approaches.

## Results

### Non-participation and rare variant burden

In 377,861 participants who were genetically similar to the 1KGP European superpopulation, we observed significant associations between non-participation in cognitive testing and greater burden of damaging RCVs and rare CNVs (Figure 2; Supplementary Table 5). The strongest effects were seen for rare NPD-associated CNVs, which were associated with higher odds of non-participation in cognitive testing (ORs between 1.42 – 2.21; Figure 2; Supplementary Table 5). Damaging RCVs in LoFi and in NPD-associated genes were also associated with non-participation in cognitive testing, but with weaker effects than those observed for CNVs (Figure 2; Supplementary Table 5). All three rare variant classes had largest effects on the strongest measure of non-participation (not attempting baseline cognitive testing), and smaller effects on the weaker measures of non-participation (not completing <u>any</u> and not completing <u>all</u> online cognitive tests) (Figure 2). Synonymous RCVs in LoFi or NPD-associated genes were not significantly associated with non-participation (Supplementary Table 5).

**Figure 2.**
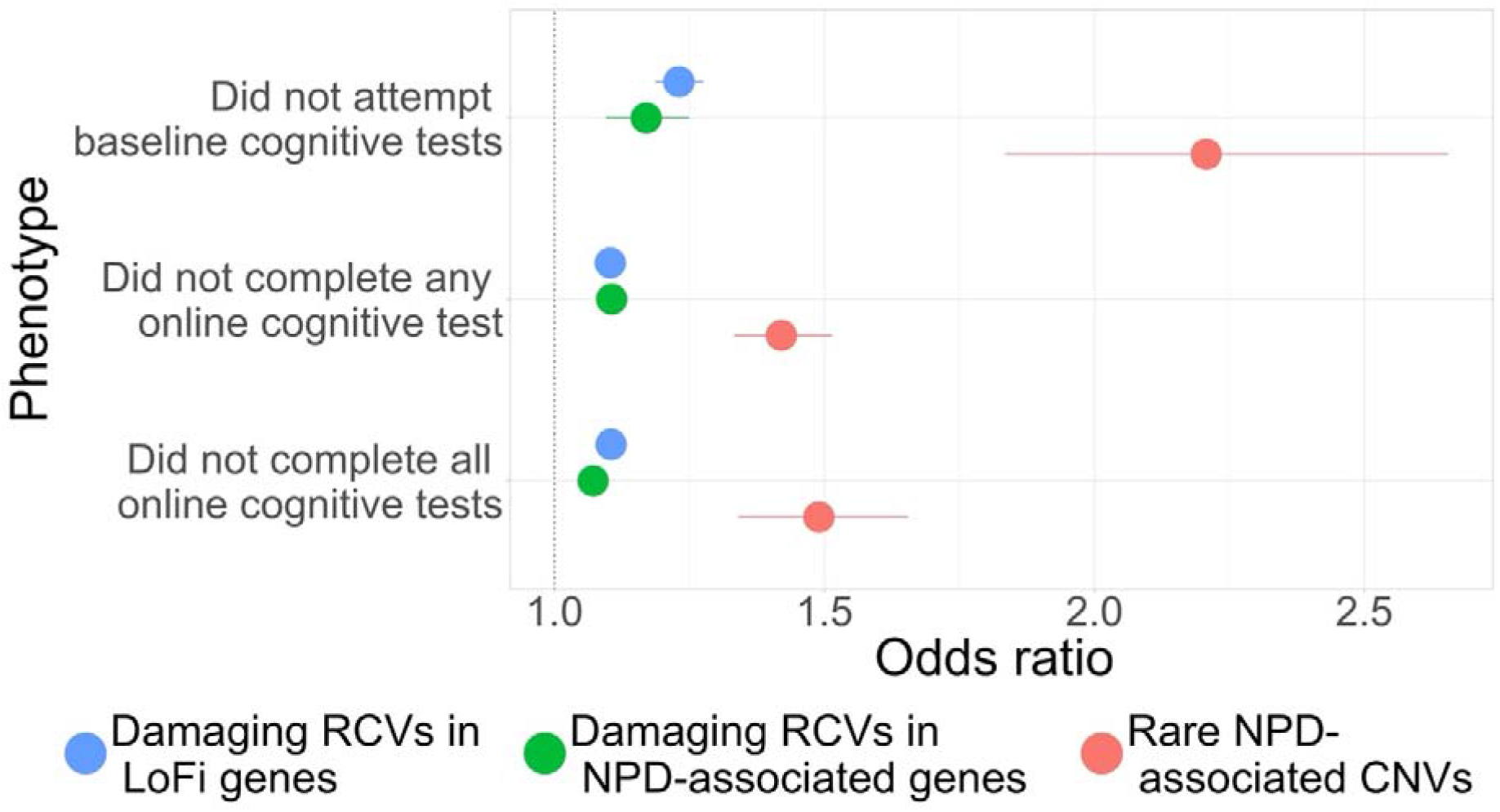
Association of non-participation in cognitive testing with rare variant burden in participants of 1KGP-EUR-like genetic ancestries. RCVs = rare coding variants; LoFi = loss-of-function intolerant; NPD = neurodevelopmental and psychiatric disorder; CNVs = copy number variants. Error bars display 95% confidence intervals.

In 8,724 participants of 1KGP-SAS-like genetic ancestries, we observed no significant associations of any class of variant with any participation phenotype, possibly reflecting limited statistical power due to the smaller sample size (Supplementary Table 6). Sample sizes of all other 1KGP-like genetic ancestries were too small (< 5,000, see Supplementary Table 1) to accurately examine the impact of rare variants on participation. Therefore, all analyses presented hereafter are based only on participants that are genetically similar to the 1KGP European superpopulation.

### Broader participation phenotypes

Similar results reflecting an association of non-participation with greater burden of damaging RCVs and rare CNVs were observed for broader participation phenotypes, including loss to follow-up, no email access, and not completing an online mental health questionnaire (Supplementary Table 7; Supplementary Figure 1). These findings suggest the observed effect of damaging rare variants on participation is not specific to participation in cognitive testing.

### Controlling for invitation, neurodevelopmental and psychiatric disorder diagnoses, and cognitive ability

In analyses covarying for invitation (via email) to online cognitive testing, all associations between rare variants and non-participation were significantly weaker (z-test p values < 0.05) compared to analyses that did not include this covariate (Supplementary Figure 2; Supplementary Table 8). While covarying for invitation attenuated the effects of rare variants on non-participation, all but one association that was significant without this covariate remained significant when controlling for invitation (Supplementary Figure 2; Supplementary Table 8). This, taken alongside the finding that rare variants are associated with not having email access (Supplementary Table 7), suggests that these variants influence both a participant’s likelihood of being invited to participate in cognitive testing, and whether they do participate given this opportunity.

Associations between damaging RCVs and rare CNVs and non-participation in cognitive testing were not significantly attenuated when adjusting for NPD diagnosis (Supplementary Figure 3; Supplementary Table 9).

Given the known associations between damaging RCVs and rare CNVs and lower cognitive ability, we examined whether cognitive ability itself mediated the effects of these variants on non-participation in cognitive testing. No associations were significantly attenuated when adjusting for RT (Supplementary Figure 4; Supplementary Table 10). Covarying for FI significantly attenuated the associations between damaging RCVs in LoFi genes and participation in online cognitive testing, as well as the association of NPD-associated CNVs with not completing <u>any</u> online cognitive test. In addition, three of the six associations that were significant in the main models were no longer significant after adjusting for FI (Supplementary Figure 5; Supplementary Table 11).

### Impact of non-participation on rare variant associations with cognitive ability

We next examined whether participation bias influenced associations between rare variants and cognitive ability in UKBB. We stratified individuals based on whether or not they had completed <u>all</u> online cognitive testing and estimated associations between damaging RCVs and CNVs and cognitive ability separately in those who completed <u>all</u> online cognitive testing and in those who did not. We then compared associations between these groups using z-tests.

Rare variants were found to have weaker effects on RT in UKBB participants who completed <u>all</u> online cognitive testing compared with those who did not (Figure 3, Supplementary Table 12). The differences in effect sizes between participation groups were significant for both classes of RCVs, but not for NPD-associated CNVs (Figure 3, Supplementary Table 12).

**Figure 3.**
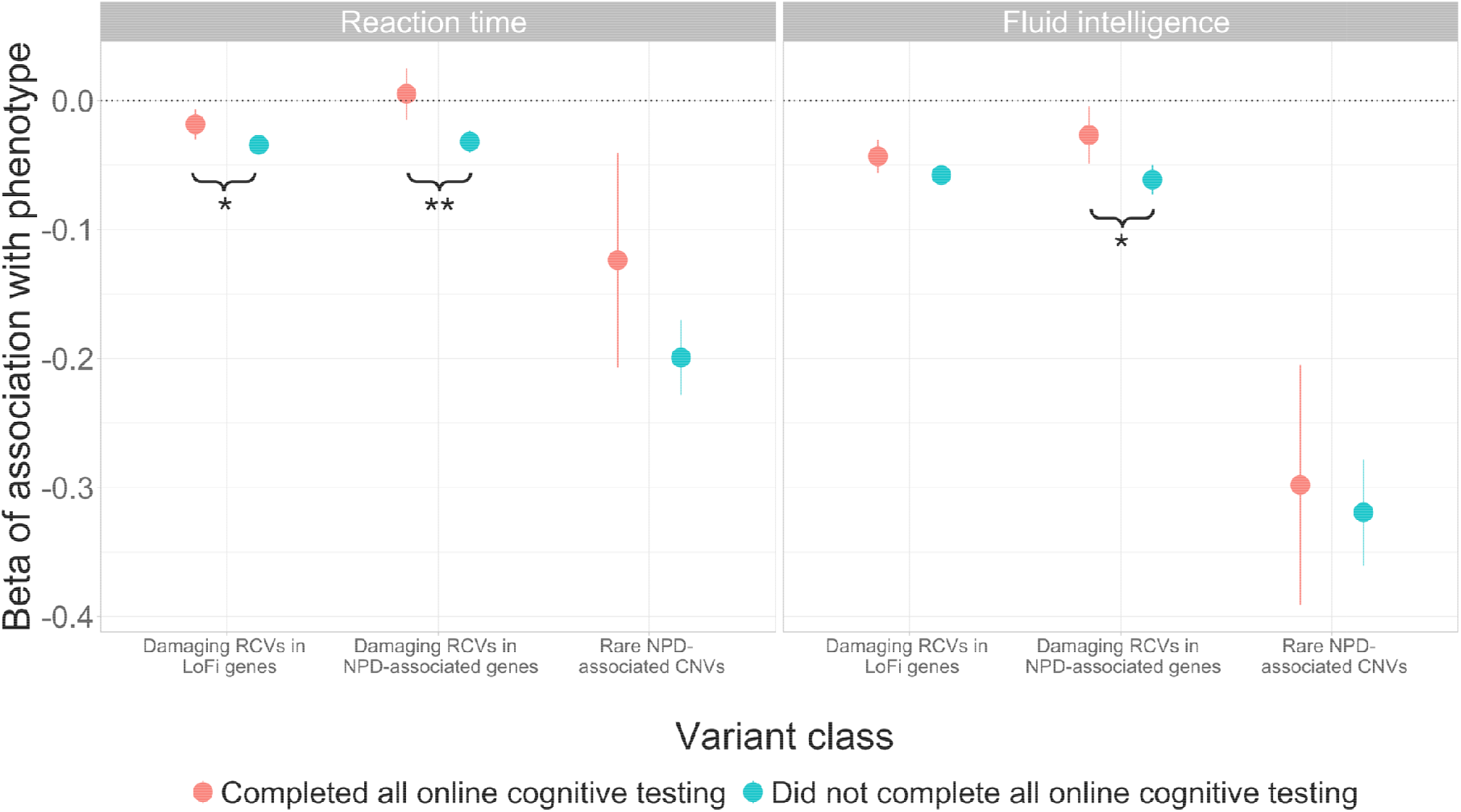
Associations of cognitive measures (reaction time and fluid intelligence) with damaging RCV and rare CNV burden in participants of 1KGP-EUR-like genetic ancestries who completed <u>all</u> online cognitive testing (red) compared to those who did not (blue). Error bars represent 95% confidence intervals. Significant results from z-tests comparing these associations are presented using braces and stars to represent significance: * p < 0.0083 (Bonferroni-corrected significance threshold); ** p < 0.001.

A similar pattern was observed for FI, with all associations being weaker in those who completed <u>all</u> online cognitive tests than in those who did not (Figure 3, Supplementary Table 12). However, for this measure of cognition, the difference between participation groups was statistically significant only for damaging RCVs in NPD-associated genes, and not for the other two classes of rare variants (Figure 3, Supplementary Table 12).

To support these findings, we ran a single model across all participants per cognitive measure and included an interaction term between rare variant burden and completion of <u>all</u> online cognitive tests. The pattern of interaction effects was consistent with findings of the stratified analyses; interaction terms indicated that associations between cognitive ability and rare variants differed by participation status (Supplementary Table 13), and damaging RCVs and rare CNVs had weaker associations with cognition in people who completed <u>all</u> online cognitive tests than those who did not.

#### Reweighting

We used inverse probability weighting to reweight participants based on their estimated probability of completing <u>all</u> online cognitive testing, as derived from demographic characteristics (described in Supplementary Table 4 and Supplementary Materials 1.3), aiming to reduce the impact of participation bias on rare variant-phenotype associations.

In a logistic regression model predicting completion of <u>all</u> online cognitive tests in 392,812 individuals with complete weighting data, all of the demographic predictors (Supplementary Table 4) were significantly associated with participation. Higher educational attainment showed the strongest positive association with completing <u>all</u> online cognitive tests, whereas living in rented accommodation, greater deprivation, self-reported non-White ethnicity, and long-term sickness were the strongest predictors of not completing <u>all</u> online cognitive tests. We derived stabilised inverse probability weights by scaling the inverse probabilities by the overall participation rate, and these weights (with outliers capped at 4SDs from the mean) ranged from 0.44 to 11.7.

Applying these weights had limited impact on the pattern of associations between rare variants and cognitive measures (Supplementary Table 14; Figure 4). Most effect sizes (9 of 12) increased in magnitude after weights were applied, and all but one had more significant p-values after reweighting. However, all three z-tests showing significant differences between participation groups in the unweighted analysis remained significant after weighting (Supplementary Table 14; Figure 4), suggesting the reweighting approach did not control for the effects of participation bias on observed rare variant-cognition associations.

**Figure 4.**
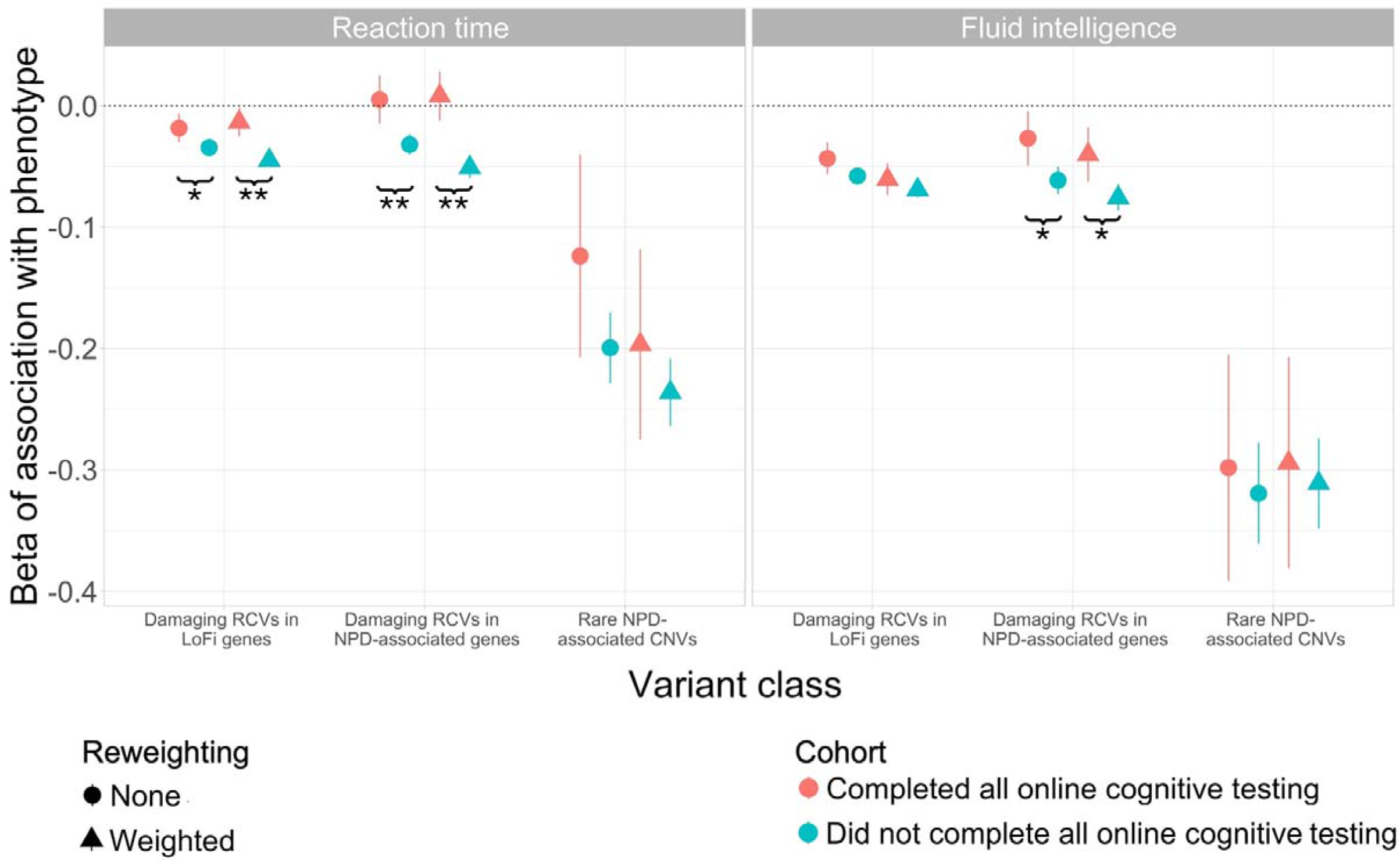
Associations of cognitive measures (reaction time and fluid intelligence) with damaging RCV and rare CNV burden in participants of 1KGP-EUR-like genetic ancestries who completed all online cognitive testing (red) compared to those who did not (blue), in the original model (circles) and using our derived weightings (triangles). Error bars represent 95% confidence intervals. Significant results from z-tests comparing these associations are presented using braces and stars to represent significance: * p <. 0.0083 (Bonferroni-corrected significance threshold); ** p < 0.001.

## Discussion

In this study, we show that individuals with lower levels of participation in optional components of UKBB have a higher burden of rare damaging RCVs in constrained and NPD-associated genes, and of NPD-associated CNVs. These findings indicate that participation in optional study components is a non-random and genetically influenced phenotype. We then demonstrate that participation bias has important implications for the generalisability of genetic analyses relying on optional study components; for example, the association between damaging RCVs in LoFi genes and RT was significantly stronger in those who did not complete <u>all</u> online cognitive tests compared to those who did, and the association between damaging RCVs in NPD-associated genes and RT was evident only in those who did not complete all online cognitive tests. This indicates that participation bias can significantly weaken, and in some cases fully attenuate, rare variant-phenotype associations. Taken together, our results highlight the need to consider the effects of participation bias when performing genetic studies in population-level cohorts. They also emphasise the need for population-level cohorts to make efforts to support and enable ongoing participation in order to avoid underrepresentation of participants with certain characteristics (e.g. lower cognitive ability, lower educational attainment), as this may lead to systematic underestimation of the effects of rare variants.

To our knowledge, this study is the first to examine participation biases across rare variant classes (damaging RCVs and NPD-associated CNVs). We observed strong associations between rare CNV burden and non-participation in cognitive testing, with carriers of NPD-associated CNVs being more than twice as likely not to attempt baseline cognitive testing compared to non-carriers (OR = 2.21). Damaging RCVs in LoFi and NPD-associated genes were also significantly associated with non-participation in cognitive testing, but with smaller effects than CNVs (ORs between 1.07 and 1.23). Previous work has shown a negative effect of a stricter definition of loss-of-function variants in autism-associated genes on response to questionnaire-based measures and participation in imaging assessments^46^. We now broaden and extend this work by demonstrating impacts of both PTVs and deleterious missense variants, and of CNVs, on participation across cognitive and non-cognitive components of UKBB. We provide novel evidence that associations of RCVs and CNVs were not fully explained by differences in opportunity to participate (e.g. invitation to online cognitive testing) or by NPD diagnoses. Importantly, unlike previous work, we also examine the consequences of participation bias for rare variant-phenotype associations.

The observed associations of RCVs in NPD-associated genes with non-participation in this study parallel previous findings for common variants; common alleles implicated in schizophrenia have been associated with non-participation over time in ALSPAC, another population-level cohort^3^. This implies that individuals with higher genetic liability for NPDs (from both common and rare variants) are underrepresented among participants who remain engaged in population-level cohorts over time. This underrepresentation reduces the generalisability of findings from analyses of NPD-associated genetic factors and/or phenotypes (e.g. cognitive ability) to less biased or clinical populations.

We examined whether the effect of rare variants on participation was specific to cognitive testing or extended to non-cognitive participation by examining broader participation phenotypes. Damaging RCV and CNV burden was associated with non-participation in non-cognitive components of UKBB (including loss to follow-up, no email access, and non-completion of the online mental health questionnaire), suggesting a broad impact of these rare variants on participation that extends beyond cognitive testing. One broad participation measure examined was email access, as only those with email access were invited to participate in online cognitive testing in UKBB. We observed associations between carrying damaging RCVs or rare CNVs and not having email access, mirroring results from studies of common variation in this cohort^1^. Overall, these findings suggest that rare and common variants impact not only participation given invitation, but also the likelihood of being offered opportunities to participate, highlighting the need for more accessible methods of invitation to follow-ups in UKBB and similar cohorts.

We hypothesised that the known association between damaging RCVs and rare CNVs and cognitive ability^7–9,11^ may mediate the associations we observed between non-participation and rare variant burden, as those with higher cognition may be more likely to complete optional cognitive testing, and less likely to carry damaging rare variants. We tested this by adjusting for cognitive ability in our analyses of non-participation and rare variant burden. Adjusting for RT did not significantly impact any associations of damaging RCVs and rare CNVs with non-participation in online cognitive testing. Conversely, three of six significant associations became non-significant after adjusting for FI score, suggesting that cognitive ability itself (or factors relating to it) may partially mediate associations between non-participation and damaging RCVs and rare CNVs. However, the FI analysis was restricted to individuals who completed the FI test at baseline assessment (34% of the total sample) and is therefore itself subject to bias.

Although our analyses focused only on participation in optional components of UKBB, similar biases may also impact initial participation in the study. Previous work has shown UKBB participants are generally healthier, less socioeconomically deprived and have higher levels of education than the general population^19,47^. It is therefore feasible that individuals with lower cognitive ability (or other factors associated with rare variant burden) are underrepresented both at initial recruitment and in follow-up assessments. If this is the case, estimates derived from the recruited sample may already underestimate the effects of rare variants, even before the additional participation biases observed in this study further weaken associations.

A key strength of this study is the use of large-scale high-quality genetic data, combined with detailed measures of participation across multiple study components, which allowed us to assess the influence of damaging RCVs and CNVs on participation, and the impact of participation bias on rare variant-phenotype associations. However, despite the large overall sample size, some analyses were underpowered, particularly those involving non-1KGP-EUR-like genetic ancestries. Future work in more diverse and representative samples will be essential to fully characterise the relationship between rare variants and participation.

The second part of this study examined the impact of non-participation on rare variant-phenotype associations, which, to our knowledge, has not yet been investigated in UKBB or similar cohorts. We found that associations of damaging RCVs and CNVs with cognitive ability were significantly weakened, and in some cases no longer detectable, when analyses were restricted to subsets of UKBB with higher participation (i.e. those who completed <u>all</u> online cognitive tests). This indicates that participation bias reduces power to detect rare-variant phenotype associations, increasing the risk of false negative findings.

We were unable to directly examine the possibility of participation bias inducing false positive findings, but we acknowledge that this is a possibility in the context of collider bias. Collider bias has the potential to arise if both the burden of rare variants (the exposure) and cognitive ability (the outcome) independently influence participation (the collider), and conditioning on this collider could induce or distort associations between rare variant burden and cognition. However, given that higher rare variant burden and lower cognitive ability were both associated with reduced participation in this study, we expect conditioning on participation would reduce power to identify true associations, rather than induce false associations between rare variant burden and cognitive ability.

Overall, our findings highlight that where possible, researchers should prioritise the use of phenotypes with minimal missingness to reduce the impact of participation bias on their results, however, this is not always feasible. We attempted to mitigate the impact of participation bias on rare variant-phenotype associations using inverse probability weighting, with weights derived from demographic characteristics, however it had limited impacts. Whilst it strengthened effect sizes and increased the significance of most associations between rare variants and cognitive ability, differences in rare variant-cognition associations between participation groups remained after reweighting. This suggests that participation in optional components of UKBB cannot be fully explained by demographic factors alone, likely reflecting the influence of unmeasured or poorly captured factors (e.g. cognitive ability, health, or genetic liability) that may continue to influence both participation and genetic association estimates and cannot be fully addressed through reweighting. Other studies have suggested alternative ways to mitigate the impacts of this bias; for example, recent work demonstrated that imputing FI scores across the whole UKBB cohort can reduce ascertainment bias, and increase power for genetic association analyses^48^. When analysing phenotypes with substantial missingness, where reweighting, imputation, or other mitigation strategies are not possible, participation bias should be carefully considered. Researchers should be mindful that restricting analyses to certain phenotypes limits the sample to individuals who tend to carry fewer damaging RCVs and rare CNVs, and whose variants may have (or may appear to have due to bias) weaker phenotypic effects.

In summary, our findings show that individuals with greater participation in optional components of UKBB carry fewer damaging RCVs and CNVs, and that damaging RCV and rare CNV burden are more weakly associated with cognitive ability in individuals with higher levels of participation than in those with lower levels of participation. This suggests that participants who engage in optional components of UKBB are not representative of the broader cohort. Our results extend current understanding of the impact of damaging rare variants on participation, and highlight the potential for participation bias to weaken, or even mask, rare variant-phenotype associations. These findings reinforce that non-participation is a genetically influenced and non-random phenotype, underscoring the need for careful consideration of participation-related factors in genetic studies using large-scale cohorts such as UKBB. This knowledge is important considering the growing reliance on UKBB and similar population-level datasets in genetic research.

## Acknowledgments

This work was supported by a UKRI MRC Programme Grant (MR/Y004094/1) to JTRW, ER, PH, MCOD and MJO, a Wellcome Trust Integrative Neuroscience PhD Studentship to EF (108891/B/15/Z /WT), and a UKRI Future Leaders Fellowship Grant to ER (MR/T018712/1 and MR/Y033922/1).

This research has been conducted using the UK Biobank Resource under Application Number 13310. This work uses data provided by patients and collected by the NHS as part of their care and support: Copyright © (2023), NHS England; Re-used with the permission of the NHS England and UK Biobank; All rights reserved. This research used data assets made available by National Safe Haven as part of the Data and Connectivity National Core Study, led by Health Data Research UK in partnership with the Office for National Statistics and funded by UK Research and Innovation (research which commenced between 1st October 2020 – 31st March 2021 (MC_PC_20029); 1st April 2021 -30th September 2022 (MC_PC_20058).

E.F., E.R., and J.T.R.W. conceived and designed the research. E.F. analysed the data. E.F., E.R., J.T.R.W., P.H., M.C.O.D. and M.J.O. contributed to the interpretation of the results. E.F., E.R., and J.T.R.W. wrote the manuscript, which was read, edited and approved by all authors.

## Data Access and Code Availability

This research was conducted using the UK Biobank Resource under Application Number 13310. Data from the UK Biobank is available for health-related research upon registration and application through the UK Biobank Access Management System (https://www.ukbiobank.ac.uk/enable-your-research/register). The code required to reproduce our analyses is publicly available (https://github.com/eilidhfenner/UKBB_participation_bias).

## References

1. Adams, M. J. et al. Factors associated with sharing e-mail information and mental health survey participation in large population cohorts. Int. J. Epidemiol. 49, 410–421 (2020).

2. Adams, M. J. Leveraging family relatedness to detect participation bias in genetic studies. Nat. Genet. 1–2 (2023) doi:10.1038/s41588-023-01440-9.

3. Martin, J. et al. Association of Genetic Risk for Schizophrenia With Nonparticipation Over Time in a Population-Based Cohort Study. Am. J. Epidemiol. 183, 1149–1158 (2016).

4. Benonisdottir, S. & Kong, A. Studying the genetics of participation using footprints left on the ascertained genotypes. Nat. Genet. 1–8 (2023) doi:10.1038/s41588-023-01439-2.

5. Tyrrell, J. et al. Genetic predictors of participation in optional components of UK Biobank. Nat. Commun. 12, 886 (2021).

6. Mignogna, G. et al. Patterns of item nonresponse behaviour to survey questionnaires are systematic and associated with genetic loci. *Nat*. Hum. Behav. 1–17 (2023) doi:10.1038/s41562-023-01632-7.

7. Fenner, E. et al. Analysis of rare coding variants in schizophrenia-associated genes and generalised cognition in the UK Biobank. Mol. Psychiatry 1–9 (2026) doi:10.1038/s41380-026-03601-8.

8. Kingdom, R. et al. Rare genetic variants in genes and loci linked to dominant monogenic developmental disorders cause milder related phenotypes in the general population. Am. J. Hum. Genet. 109, 1308–1316 (2022).

9. Chen, C.-Y. et al. The impact of rare protein coding genetic variation on adult cognitive function. Nat. Genet. 1–12 (2023) doi:10.1038/s41588-023-01398-8.

10. Kendall, K. M. et al. Cognitive Performance Among Carriers of Pathogenic Copy Number Variants: Analysis of 152,000 UK Biobank Subjects. Biol. Psychiatry 82, 103– 110 (2017).

11. Kendall, K. M. et al. Cognitive performance and functional outcomes of carriers of pathogenic copy number variants: analysis of the UK Biobank. Br. J. Psychiatry 214, 297–304 (2019).

12. Taylor, A. E. et al. Exploring the association of genetic factors with participation in the Avon Longitudinal Study of Parents and Children. Int. J. Epidemiol. 47, 1207–1216 (2018).

13. Munafò, M. R., Tilling, K., Taylor, A. E., Evans, D. M. & Davey Smith, G. Collider scope: when selection bias can substantially influence observed associations. Int. J. Epidemiol. 47, 226–235 (2018).

14. Stamatakis, E. et al. Is Cohort Representativeness Passé? Poststratified Associations of Lifestyle Risk Factors with Mortality in the UK Biobank. Epidemiology 32, 179 (2021).

15. Bradley, V. & Nichols, T. E. Addressing selection bias in the UK Biobank neurological imaging cohort. 2022.01.13.22269266 Preprint at 10.1101/2022.01.13.22269266 (2022).

16. Keyes, K. M. & Westreich, D. UK Biobank, big data, and the consequences of non-representativeness. The Lancet 393, 1297 (2019).

17. van Alten, S., Domingue, B. W., Faul, J., Galama, T. & Marees, A. T. Correcting for volunteer bias in GWAS increases SNP effect sizes and heritability estimates. Nat. Commun. 16, 3578 (2025).

18. van Alten, S., Domingue, B. W., Faul, J., Galama, T. & Marees, A. T. Reweighting UK Biobank corrects for pervasive selection bias due to volunteering. Int. J. Epidemiol. 53, dyae054 (2024).

19. Schoeler, T. et al. Participation bias in the UK Biobank distorts genetic associations and downstream analyses. Nat. Hum. Behav. 1–12 (2023) doi:10.1038/s41562-023-01579-9.

20. Van Hout, C. V. et al. Exome sequencing and characterization of 49,960 individuals in the UK Biobank. Nature 586, 749–756 (2020).

21. Hail Team. Hail. https://github.com/hail-is/hail.

22. Lin, M. F., et al. GLnexus: joint variant calling for large cohort sequencing. bioRxiv 343970 (2018) doi:10.1101/343970.

23. Relationship Inference in KING. https://www.kingrelatedness.com/manual.shtml.

24. Manichaikul, A. et al. Robust relationship inference in genome-wide association studies. Bioinformatics 26, 2867–2873 (2010).

25. McLaren, W. et al. The Ensembl Variant Effect Predictor. Genome Biol. 17, 122 (2016).

26. Kingdom, R., Beaumont, R. N., Wood, A. R., Weedon, M. N. & Wright, C. F. Genetic modifiers of rare variants in monogenic developmental disorder loci. Nat. Genet. 56, 861–868 (2024).

27. Karczewski, K. J. et al. The mutational constraint spectrum quantified from variation in 141,456 humans. Nature 581, 434–443 (2020).

28. Karczewski, K. konradjk/loftee. (2020).

29. Ioannidis, N. M. et al. REVEL: An Ensemble Method for Predicting the Pathogenicity of Rare Missense Variants. Am. J. Hum. Genet. 99, 877–885 (2016).

30. Lek, M. et al. Analysis of protein-coding genetic variation in 60,706 humans. Nature 536, 285–291 (2016).

31. Gardner, E. J. et al. Reduced reproductive success is associated with selective constraint on human genes. Nature 603, 858–863 (2022).

32. DDG2P (version 5.1). Accessed on 08/11/2024. https://panelapp.genomicsengland.co.uk/panels/484/.

33. Thormann, A. et al. Flexible and scalable diagnostic filtering of genomic variants using G2P with Ensembl VEP. Nat. Commun. 10, 2373 (2019).

34. Singh, T. et al. Rare coding variants in ten genes confer substantial risk for schizophrenia. Nature 1–9 (2022) doi:10.1038/s41586-022-04556-w.

35. Trubetskoy, V. et al. Mapping genomic loci implicates genes and synaptic biology in schizophrenia. Nature 604, 502–508 (2022).

36. Stefansson, H. et al. CNVs conferring risk of autism or schizophrenia affect cognition in controls. Nature 505, 361–366 (2014).

37. Huddart, R. et al. Standardized Biogeographic Grouping System for Annotating Populations in Pharmacogenetic Research. Clin. Pharmacol. Ther. 105, 1256–1262 (2019).

38. Legge, S. E. et al. A genome-wide association study in individuals of African ancestry reveals the importance of the Duffy-null genotype in the assessment of clozapine-related neutropenia. Mol. Psychiatry 24, 328–337 (2019).

39. Smart, S. E. et al. The Role of SLC39A8.p.(Ala391Thr) in Schizophrenia Symptom Severity and Cognitive Ability: Cross-Sectional Studies of Schizophrenia and the General UK Population. Am. J. Med. Genet. B Neuropsychiatr. Genet. 198, 135–146 (2025).

40. Auton, A. et al. A global reference for human genetic variation. Nature 526, 68–74 (2015).

41. Yang, N. et al. Examination of ancestry and ethnic affiliation using highly informative diallelic DNA markers: application to diverse and admixed populations and implications for clinical epidemiology and forensic medicine. Hum. Genet. 118, 382–392 (2005).

42. Huang, Q. Q. et al. Examining the role of common variants in rare neurodevelopmental conditions. Nature 636, 404–411 (2024).

43. Kosmidis, I. brglm: Bias Reduction in Binomial-Response Generalized Linear Models. (2021).

44. Kosmidis, I. & Firth, D. Jeffreys-prior penalty, finiteness and shrinkage in binomial-response generalized linear models. Biometrika 108, 71–82 (2021).

45. Green, E. et al. Exploring patterns of response across the lifespan: the Cambridge Centre for Ageing and Neuroscience (Cam-CAN) study. BMC Public Health 18, 760 (2018).

46. Rolland, T. et al. Phenotypic effects of genetic variants associated with autism. Nat. Med. 1–10 (2023) doi:10.1038/s41591-023-02408-2.

47. Fry, A. et al. Comparison of Sociodemographic and Health-Related Characteristics of UK Biobank Participants With Those of the General Population. Am. J. Epidemiol. 186, 1026–1034 (2017).

48. van den Berg, D. M., et al. Imputation of fluid intelligence scores reduces ascertainment bias and increases power for analyses of common and rare variants. Nat. Genet. 1–10 (2026) doi:10.1038/s41588-026-02787-5.

