## Supplementary Materials and Figures 1-5 for "Rare genetic variants influence participation in cognitive components of the UK Biobank"

**Author list and affiliations:**

Eilidh Fenner^1^, Peter Holmans^1^, Michael C O’Donovan^1^, Michael J Owen^1^, Elliott Rees^1+*^, James T R Walters^1+*^

1. Centre for Neuropsychiatric Genetics and Genomics, Department of Psychological Medicine and Clinical Neurosciences, Cardiff University, Cardiff, UK

^+^ these authors contributed equally to this work. *Corresponding authors.

### 1 Supplementary Materials

#### 1.1 Broad participation phenotypes

We investigated three broad and non-cognitive participation-related phenotypes to examine the broader role of rare variants on participation. These phenotypes are described below:

**Lost to follow-up:** Participants have been invited to participate in various follow-up measures since baseline assessment. 119,871 participants (30.1%) have had no contact with UKBB since baseline assessment (defined using UKBB field 20145) and are defined as being lost to follow-up.

**No email access:** At the baseline assessment centre and follow up assessments, participants were asked to provide an email address for recontact. 160,379 participants (40.3%) have not provided a working email address (UKBB field 20005) and are classified as having no email access.

**Did not complete mental health questionnaire:** UKBB participants were invited via email to complete an online mental health questionnaire. 271,722 participants (68.3%) did not complete this questionnaire (defined using UKBB field 20400).

#### 1.2 Phenotypes reflecting cognitive ability

We selected the reaction time (RT) and fluid intelligence (FI) tests as measures of cognitive ability, described below:

**RT**: This test reflects processing speed and is similar to a game of ‘snap’ played across 12 rounds. The participant was shown two cards at a time and asked to press a button if they matched. We used the mean time taken to correctly identify matches as our measure of RT (UKBB Field 20023), and thus a higher value on this measure reflects a longer reaction time and therefore worse cognition. This measure was non-normally distributed and was therefore log-transformed and then converted to a z-score. It was then multiplied by minus 1 so a higher score reflected better performance. Individuals with outlying scores (outside of +/- 4 SDs of the mean) were excluded from this analysis.

**FI:** This test reflects reasoning ability and was completed across baseline, later assessment centres, and as part of the online cognitive test batteries. In this test, participants were presented with 13 multiple-choice questions designed to measure their problem solving, logic, and reasoning capacity, independent of acquired knowledge. They were given 2 minutes to answer as many questions as possible, and the questions got progressively harder. We used the number of correct answers as our measure of FI, and thus a higher score on this measure reflects more questions answered correctly, and better cognition. We used test scores from assessment centres and online tests to maximise our sample size with FI (Supplementary Table 3). Where participants had completed the FI test multiple times, we selected the FI score from the first time they completed this test (e.g. if a participant had an FI score from baseline and from a later online cognitive test battery, we used the score from the baseline assessment). This measure was normally distributed. We converted this measure to a z-score, and there were no outliers outside of +/-4SDs of the mean.

#### 1.3 Demographic phenotypes for reweighting

Our reweighting phenotypes were selected as phenotypes available for > 99% of the cohort at baseline assessment, which have been previously used in reweighting studies of the UKBB (Supplementary Table 4). For the reweighting analysis, we kept only individuals who were alive at the time of data collection (i.e. alive when invited to both online cognitive tests) and who had non-missing data across all of these phenotypes (n = 392,812). Our reweighting variables were coded as described below. Many categorical variables were collapsed into fewer categories to reduce sparsity and ensure more stable weighting.

**Age** (UKBB Field 21022): Derived by the UKBB using the date of birth and date of attending the initial assessment centre, truncated to a whole year. We standardised age across the cohort. No participants had missing data for this variable.

**Breastfed as a baby** (UKBB Field 1677): Participants were asked via a touchscreen device “*Were you breastfed as a baby?”.* Response options were: “*Yes*” (n = 220,023); “*No*” (n = 84,294); and “*Do not know*” and “*Prefer not to answer*” (combined into one category (n = 93,385). 447 participants had missing data for this variable.

**Country of birth** (UKBB Field 1647): Participants were asked where they were born and were given the following response options*: “England”; “Wales”; “Scotland”; “Northern Ireland”; “Republic of Ireland”; “Elsewhere”; “Do not know”;* and “*Prefer not to answer*”. These were collapsed into “England” (n = 305,786), “Other UK and the Republic of Ireland” (n = 56,542), “Elsewhere” (n = 34,630), and “Unknown” (n = 744). 447 participants had missing data for this variable.

**Education** (UKBB Field 6138): Participants were asked about their qualifications. We selected their highest qualification and collapsed these ordinally as follows: Unknown, including those who responded only “*Prefer not to answer*” (n = 4,342); Low, including those who responded “*None of the above*” (n = 65,607); Other, including individuals with “*Other professional qualifications*” and “*NVQ/HND/HNC*” (n = 46,384); Secondary education, including individuals with “*CSEs*” and “*O levels/GCSEs*” (n = 104,079); A levels, including individuals with “*A levels/AS levels*” (n = 44,837); and Post-Secondary Education, including individuals with “*College / University Degree*” (n = 132,395). 505 participants had missing data for this variable.

**Employment status** (UKBB Field 6142): Participants were asked about their current employment status. They could select multiple options, which we categorised hierarchically as follows: “*In paid employment or self-employed*” (n = 228,589); Other, included those who said they were a “*Full or part-time student*”, “*Looking after home and/or family*” or “*Doing unpaid or voluntary work*” (n = 24,400); “*Unable to work due to sickness or disability*” (n = 14,136); “*Unemployed*” (n = 6,499); “*Retired*” (n = 120,180); and Unknown, including those who responded “*None of the above*” or “*Prefer not to answer*” (n = 3,903). 442 participants had missing data for this variable.

**Housing tenure** (UKBB Field 680): Participants were asked “*Do you own or rent the accommodation that you live in?*”. We collapsed their responses into the following categories: Own, including those who owned their property, either outright or with a mortgage, or who had shared ownership of a property (n = 351,698); Rent, including those who rent from local authority, council, housing association, private landlord, or letting agency (n = 37,065); Other, including those who live in accommodation rent free or who none of the options applied to (n = 4,945); and Unknown, including those who preferred not to answer (n = 2,901). 1540 participants had missing data for this variable.

**Relationship status** (UKBB field 6141): This was derived as whether a participant was co-habiting with a “*Husband, wife or partner*” as the UKBB did not directly ask participants about their relationship status at baseline assessment. 288,575 participants reported living with a partner and thus were coded as being in a relationship. 1,540 participants had missing data for this variable.

**Self-reported ethnic background** (UKBB Field 21000): Participants were asked “*What is your ethnic group?*” and we used the broad categories assigned by the UKBB: “*White*” (n = 373,228); “*Mixed*”, which we combined with the “*Other ethnic group*” category (n = 4,845); “*Asian or Asian British*” (n = 9,055); “*Black or Black British*” (n = 7,647); “*Chinese*” (n = 1,399); and we combined “*Do not know*” and “*Prefer not to answer*” into an Unknown category (n = 1,491). 484 participants had missing data for this variable.

**Sex** (UKBB field 31): Acquired from central registry at recruitment, and in some cases updated by the participant. This field thus contains a mixture of NHS recorded sex and self-reported sex. No participants had missing data for this variable.

**Townsend deprivation index** (UKBB field 22189): A score (assigned by the UKBB) corresponding to a participants postcode area which was calculated immediately prior to the participant joining the UKBB and was based on the preceding national census output areas. No participants had missing data for this variable.

#### 1.4 Additional covariates

To examine whether findings of our primary analyses were driven by any specific factors, we added additional covariates to our model testing for association between our participation phenotypes and rare variants (RVs). These additional covariates included:

**Invitation to cognitive testing:** Derived from UKBB Field 22017. Participants were invited to participate in online cognitive testing via email, and as not all participants had provided an email address, some people were not invited to participate in these online tests.

**Neuropsychiatric disorder diagnoses:** These were defined as a record (from primary care data, hospital inpatient data, death register records, or self-report) of schizophrenia or related disorders (ICD10 codes F20-29), intellectual disability (ICD10 codes F70-79), ASD (ICD10 code F84), or epilepsy and related disorders (ICD10 codes G40 and G41) (n = 9,439).

**Cognitive ability**: We covaried for both FI (at baseline) and RT separately. These phenotypes are described in Supplementary Materials 1.2.

### 2 Supplementary Figures

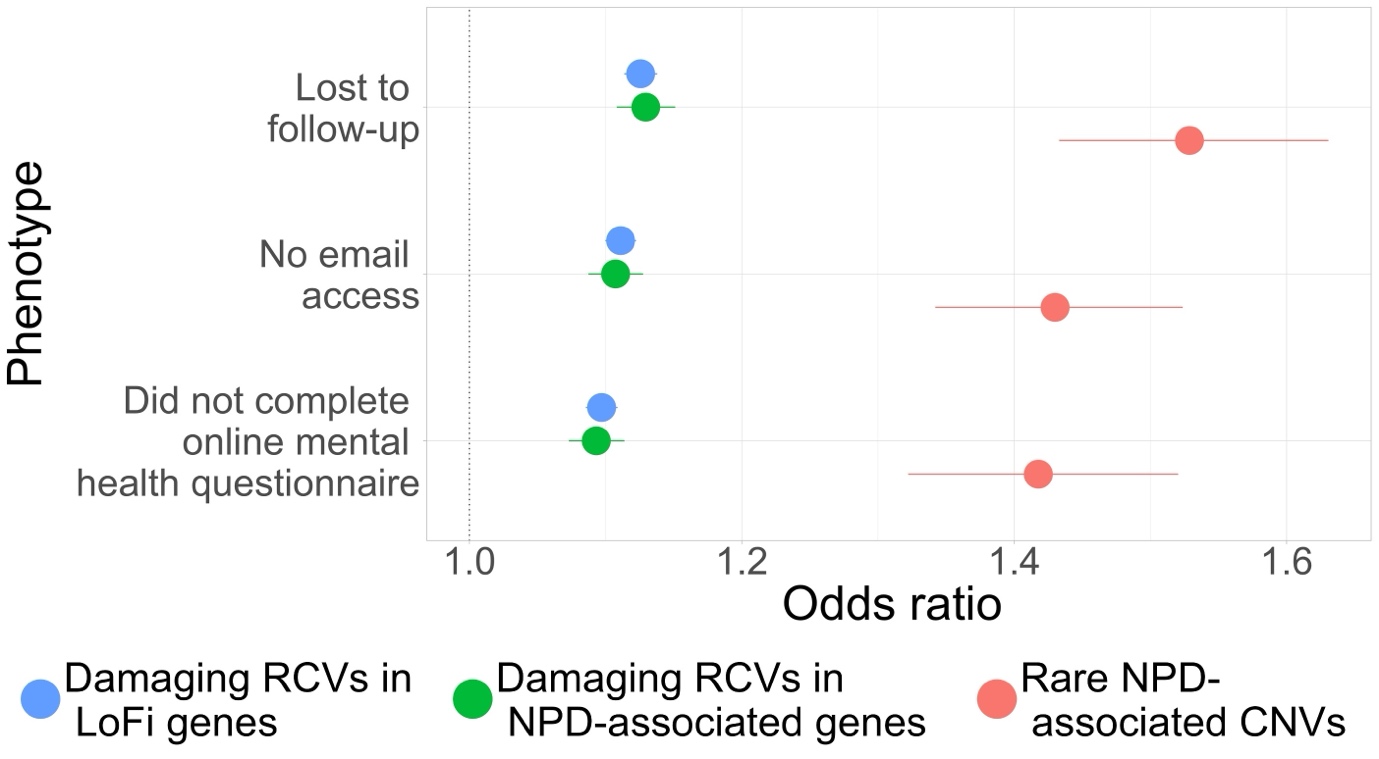

Supplementary Figure 1 - Association of broad, non-cognitive measures of non-participation with rare variant burden in participants of 1KGP-EUR-like genetic ancestries. RCVs = rare coding variants; LoFi = loss-of-function intolerant; NPD = neurodevelopmental and psychiatric disorder; CNVs = copy number variants. Error bars display 95% confidence intervals.

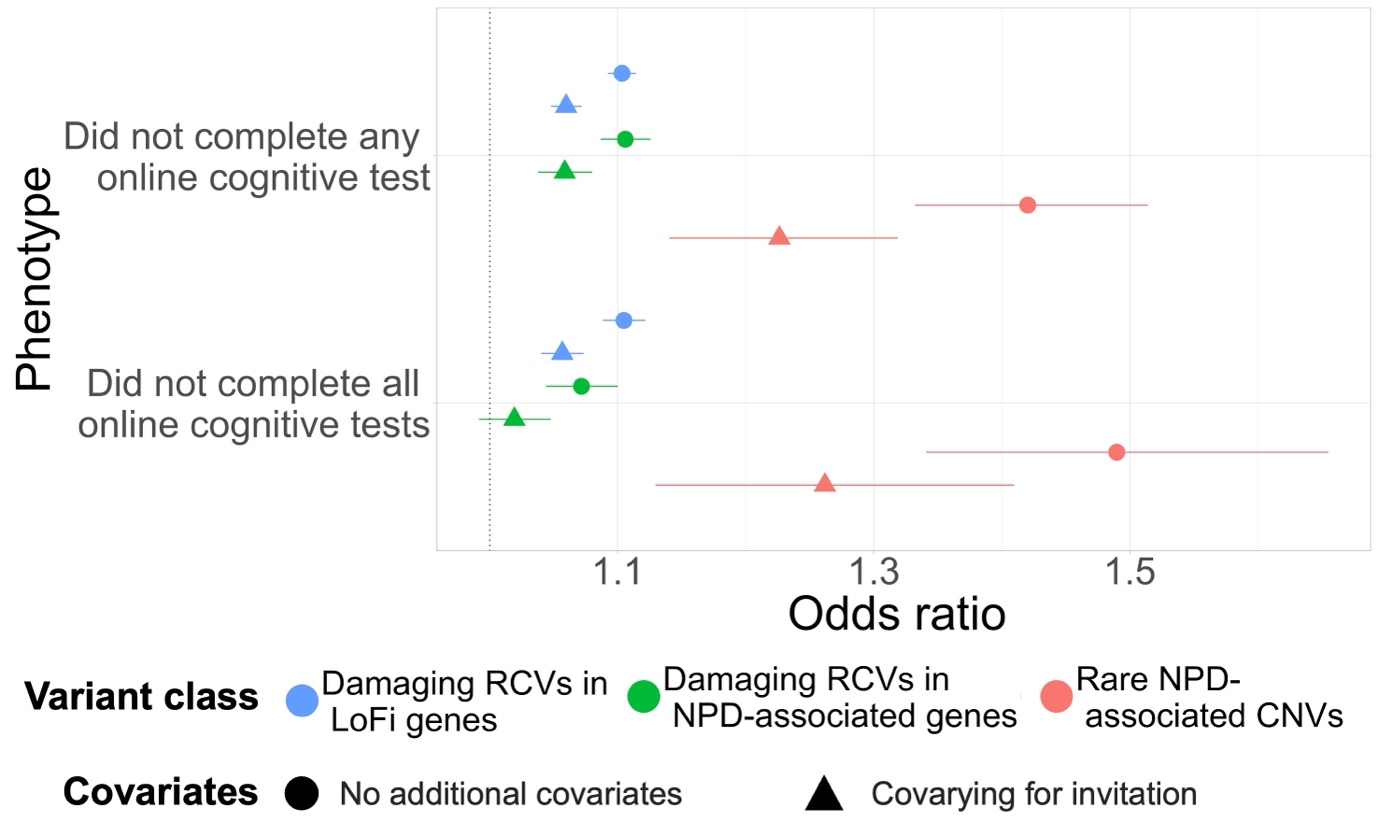

Supplementary Figure 2- Association of non-participation in cognitive testing with rare variant burden with and without covarying for invitation to online cognitive testing in participants of 1KGP-EUR-like genetic ancestries. RCVs = rare coding variants; LoFi = loss-of-function intolerant; NPD = neurodevelopmental and psychiatric disorder; CNVs = copy number variants. Error bars display 95% confidence intervals.

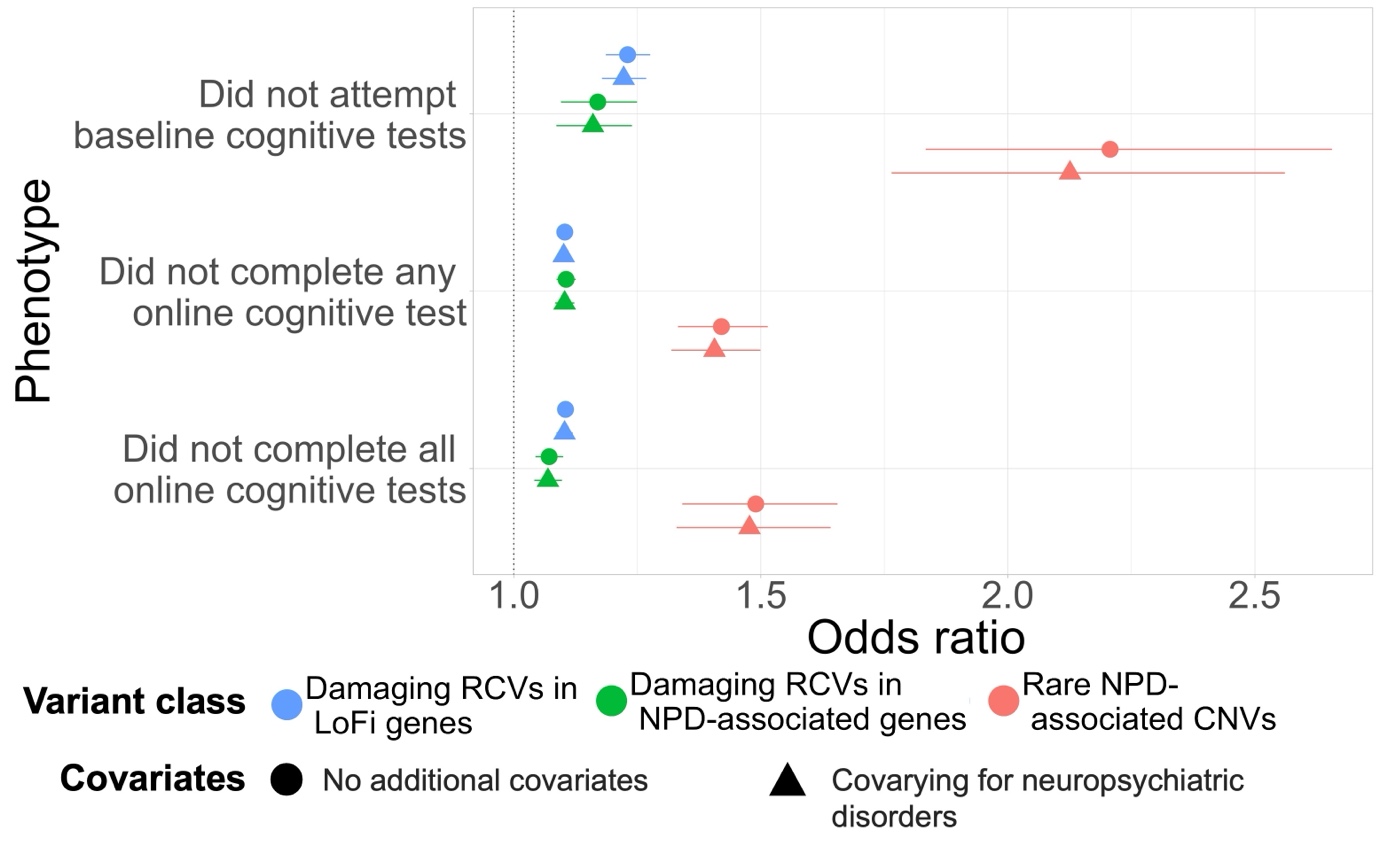

Supplementary Figure 3- Association of non-participation in cognitive testing with rare variant burden with and without covarying for neuropsychiatric disorder diagnoses in participants of 1KGP-EUR-like genetic ancestries. RCVs = rare coding variants; LoFi = loss-of-function intolerant; NPD = neurodevelopmental and psychiatric disorder; CNVs = copy number variants. Error bars display 95% confidence intervals.

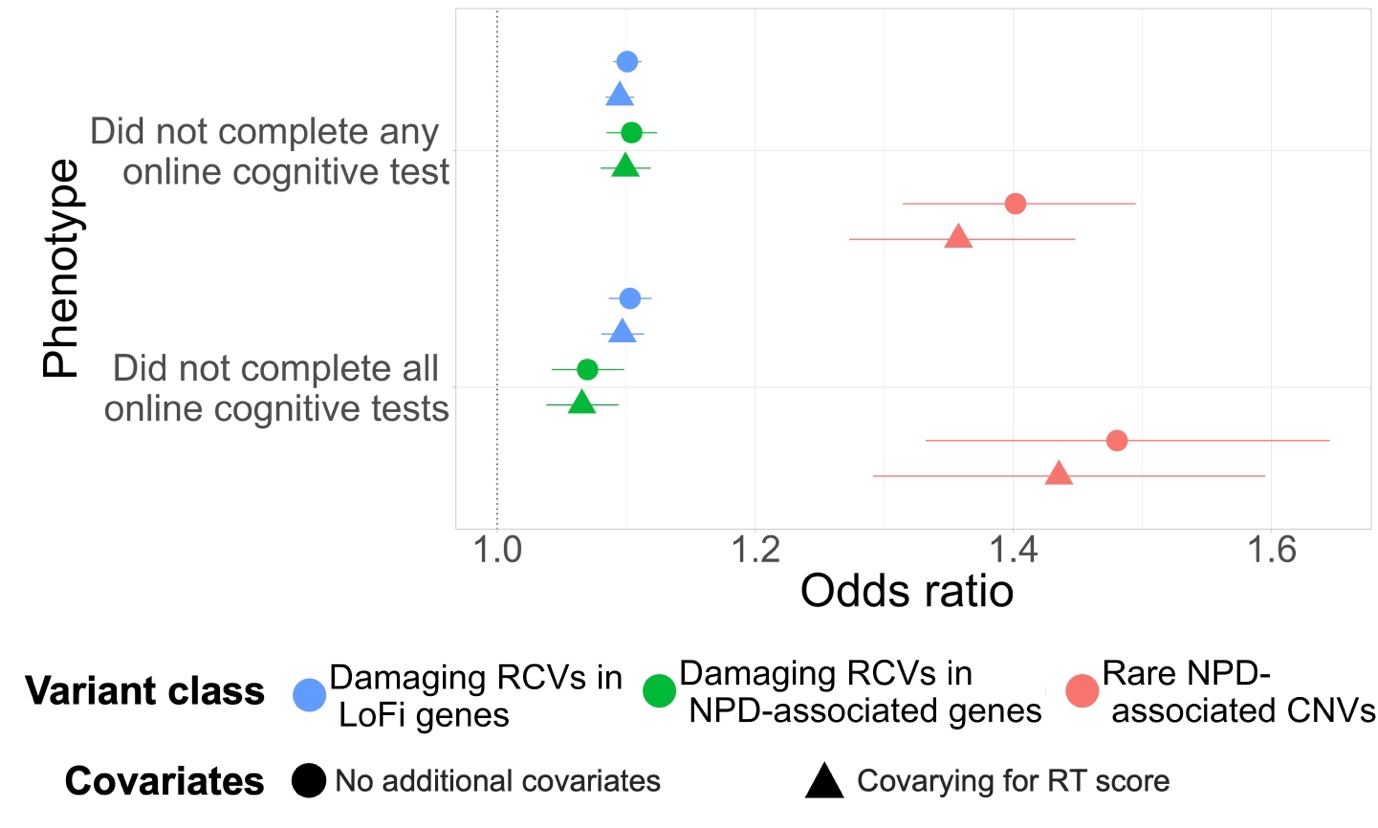

Supplementary Figure 4 - Association of non-participation in cognitive testing with rare variant burden in the subset of the 1KGP-EUR-like genetic ancestries cohort with a recorded reaction time (RT) score at baseline assessment (n = 374,413), with and without a covariate for RT score. RCVs = rare coding variants; LoFi = loss-of-function intolerant; NPD = neurodevelopmental and psychiatric disorder; CNVs = copy number variants. Error bars display 95% confidence intervals.

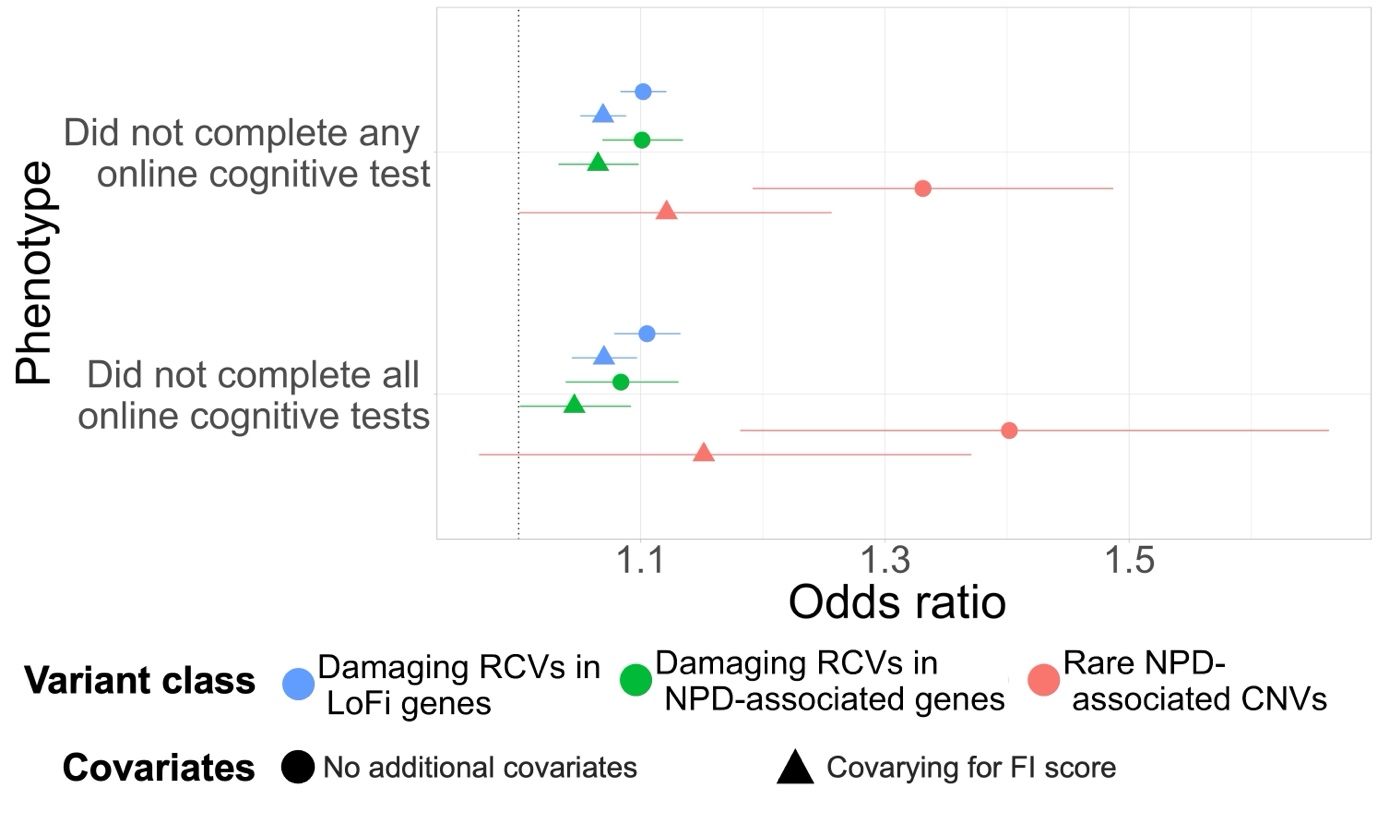

Supplementary Figure 5 - Association of non-participation in cognitive testing with rare variant burden in the subset of the 1KGP-EUR-like genetic ancestries cohort with a recorded fluid intelligence (FI) score at baseline assessment (n = 124,040), with and without a covariate for FI score. RCVs = rare coding variants; LoFi = loss-of-function intolerant; NPD = neurodevelopmental and psychiatric disorder; CNVs = copy number variants. Error bars display 95% confidence intervals.
